# Psychometric evaluation of the Severity of Violence Against Women Scale among Zambian and South African women

**DOI:** 10.1101/2025.11.13.25340160

**Authors:** Amy Zheng, Jeremy C. Kane, Saphira Munthali-Mulemba, Kristina Metz, Sristhi Sardana, Ross Greener, Sithabile Mngadi, Pertunia Manganye, Lawrence C. Long, Donald M. Thea, Laura Murray, Matthew P. Fox, Mwamba Mwenge, Anthony J. Rosellini, Sophie Pascoe

## Abstract

The Severity of Violence Against Women Scale (SVAWS) questionnaire was originally developed and validated among US women. Little is known about SVAWS’ properties in other populations and cultures. Exploratory factor analysis (EFA) was conducted among women reporting intimate partner violence (IPV) in the past year in Zambia (n=246) and South Africa (n=368). EFA indicated a three-factor solution: Simple Assault, Aggravated Assault, and Sexual Violence in South Africa and Non-Contact Violence, Physical Assault, and Sexual Violence in Zambia. While SVAWS is appropriate for assessing IPV across populations, underlying constructs may differ across cultures requiring revisions in subscale definitions to reduce bias.

**Trial Registry:** Both studies were registered on ClinicalTrials.gov: South Africa (NCT0424992) and NCT02790827 (Zambia).

## Introduction

Globally, an estimated 736 million women have experienced intimate partner violence (IPV), defined as physical, sexual, emotional, and/or psychological abuse and violence within a partnership (1), at least once in their life (2,3). IPV is associated with an increased risk of several adverse mental and physical health outcomes including depression, unplanned pregnancies, and HIV/STI transmission (2,4–9). Furthermore, IPV has economic effects of reduced productivity and increased healthcare costs (7,10–12). Reducing IPV is key to helping countries achieve the United Nations Sustainable Development Goals, in particular Goal 5, which is focused on achieving gender equality and empowering all women and girls by 2030 (13,14). Sub-Saharan Africa has the highest prevalence of IPV in the world with 43% of women reporting lifetime physical, sexual, and/or emotional IPV (15). Two fifths of women in Zambia (43%) and South Africa (40%) have reported lifetime experiences of physical and/or sexual IPV (16–18). In 2018, 28% of women in Zambia and 13% of women in South Africa reported IPV in the past 12-months (19).

Despite the abundance of evidence demonstrating the high prevalence of IPV and evidence that IPV leads to worse health and economic outcomes for women, IPV is often measured as a binary (Yes/No) outcome which does not capture frequency and severity. Prior research has demonstrated that the use of a binary measure is highly limiting and/or can lead to the incorrect conclusions (20). Specifically, an intervention may be highly effective in reducing the frequency and/or severity of IPV but not eliminate IPV (with full elimination being rare). When IPV is captured as a binary measure this would then lead to the incorrect conclusion that an intervention is ineffective when it is beneficial in reducing violence, leading to effective interventions being no longer used (20).

The Severity of Violence Against Women Scale (SVAWS) is a 46-item questionnaire that was developed to evaluate the frequency of threats and violence perpetrated by partners over the past year (21). The use of SVAWS to measure IPV in intervention studies has grown as it measures the frequency and severity of IPV and appears to capture changes in IPV after intervention (22,23). The scale was first validated among US women with an initial psychometric validation study (Marshall, 1992) concluding that the SVAWS assessed two main types (subscales) of IPV: *physically threatening acts* and *actual violence* (Murray et al., 2020; Pascoe et al., 2022). At the same time, some results of the initial validation study may be inconsistent with this conclusion. For example, eigenvalues suggested that the items measured four meaningful dimensions of violence (i.e., four factors with eigenvalues >1: *violence threats, symbolic violence, physical violence, sexual violence),* consistent with the study’s goal to assess at a minimum these four (established) types of violence (Marshall, 1992). Despite this, the authors decided to extract nine violence dimensions to create a “comprehensive and sensitive scale” (p. 107) and then used summed scores of the nine dimensions in a secondary exploratory factor analysis (EFA), which led to the final two-factor solution (21). This approach to psychometric validation deviates from gold standard recommendations (e.g., extracting many more factors than indicated by eigenvalues or parallel analysis; secondary factor analysis of summed scale scores rather than second-order EFA of individual items (Brown, 2015; Schmitt et al., 2018). Further, although the two SVAWS dimensions were found to have acceptable internal consistency, there was not additional evaluation of whether these dimensions truly measured what they were intended to (e.g., convergent and discriminant validity).

In addition to questions surrounding structural validity (how to best conceptualize the underlying constructs being measure) and convergent validity (nature of associations with other constructs/scales), little is known about SVAWS’ psychometric properties among other populations and cultures. Prior research has demonstrated that cultural beliefs and public perceptions play a key role in informing attitudes and beliefs relating to IPV (e.g., help-seeking behavior, coping mechanisms, etc.) and how IPV manifests (e.g., the types and frequency of violence) (26–32). To our knowledge, there are no published studies evaluating whether SVAWS is an appropriate tool to capture IPV in sub-Saharan Africa. Therefore, we sought to evaluate the psychometric properties of SVAWS among women in Zambia and South Africa, two populations that are diverse and distinct from women in the US and where beliefs, attitudes, and acts of violence may be differ from the US.

## Methods

### Sample

Women from Zambia (n=246) and South Africa (n=368) who reported IPV and were enrolled from May 23, 2016-December 17, 2016 in Zambia and November 11, 2021-July 19, 2023 in South Africa in a randomized control trial evaluating the effects of the Common Elements Treatment Approach (CETA), a mental health intervention, completed the SVAWS as part of their baseline measures for enrollment in the trial. Details of these two separate single-blind, parallel-assignment randomized control trials have been published elsewhere but are briefly reviewed below (22,23).

In Zambia, women were recruited from three high-density, low-socioeconomic neighborhoods in Lusaka (22,33). To be eligible for the study, women had to: 1) be at least 18 years of age, 2) be in a current relationship with a male partner, 3) speak English, Bemba, or Nyanja (the three main languages in Zambia), 4) report moderate levels of IPV within the last year (SVAWS score of 38+), and 4) have a male partner who scored 8 or more on the Alcohol Use Disorders Identification Test (e.g., an indicator of hazardous alcohol use) (22,34). Women were not eligible for the study if they: 1) were currently psychotic, 2) were currently on an unstable (e.g., altered in the last 2 months) psychiatric regimen, 3) reported a recent suicide attempt or recent suicidal ideation with specific intent, plan, or self-harm, and/or 4) had a severe developmental disorder (22).

In South Africa, women were recruited from two HIV clinics in Johannesburg. To be eligible women had to: 1) be at least 18 years of age, 2) have initiated HIV treatment; 3) have had an unsuppressed viral load (> 50 copies/mL) or had a missed or late visit within in the past year, 4) be literate and speak: English, Zulu, or SeSotho, 5) have reported any IPV on the SVAWS questionnaire and indicated that this was in the last 12 months, and 6) have their own phone which could receive text messages. Women were not eligible for the study if they: 1) were currently psychotic, 2) were currently on an unstable psychiatric regimen, 3) reported a recent a suicide attempt or recent suicidal ideation with a specific intent, plan, or self-harm, and/or 4) enrolled in any other HIV treatment intervention study (23).

The key difference between the two studies is that in Zambia a male partner had to enroll into the study as part of the eligibility criteria, while in South Africa a male partner was not required, and the women had to have initiated HIV treatment.

### Data Collection

Several questionnaires were completed at enrollment in each trial: the SVAWS, the Center for Epidemiological Studies Depression Scale (CES-D), the Harvard Trauma Questionnaire (HTQ), and the Alcohol, Smoking, and Substance Involvement Screening Test (ASSIST), all of which were administered using Audio Computer-Assisted Self Interview (ACASI). This software allows participants to read each question by themselves whilst listening to a recording of each question being read aloud using headphones. ACASI was used given the sensitive nature of IPV and the related questionnaires and has been found to reduce underreporting of sensitive topics in sub-Saharan Africa (35). Participants chose which language they wanted to complete the assessment in (South Africa: English, Zulu, or SeSotho; Zambia: English, Bemba, or Nyjanja). All translations of the surveys (e.g., SVAWS, CES-D, HTQ, and ASSIST) involved two professional translators fluent in English and the second language. After initial translation, the second translator would back translate the survey into English to ensure the non-English version retained semantic equivalency. Any differences in translation were resolved through discussion.

### The Severity of Violence Against Women Scale

The SVAWS is a 46-item questionnaire that asks women to report the frequency of violent behaviors and threats by their partner within the *past 12 months* (21). Participants indicate the frequency of each item on a Likert-scale ranging from 1 (Never) to 4 (Many Times).

### Validity Measures

The HTQ and CES-D were used as measures of convergent validity while the ASSIST was used to evaluate discriminant validity. Convergent validity was used to assess if constructs measured by SVAWS were associated with other related constructs in expected ways. In other words, the HTQ and CES-D measure related constructs of trauma exposure (i.e., IPV may be a form of trauma) and depression symptoms (recent IPV is associated with recent depression). In contrast, ASSIST was used as a comparison to evaluate discriminant validity, as IPV should be less strongly associated with lifetime substance use behaviors than with trauma or recent depression symptoms (e.g., using alcohol once at 15 years old should not be associated with recent IPV).

The HTQ consisted of two parts: 1) 17-items that evaluated lifetime exposure to traumatic events and 2) 39-items that evaluated post-traumatic symptoms in the past week (36). The second part of the HTQ assessing post-traumatic symptoms was used to test concurrent validity (Mollica et al., 1992). Symptoms are assessed using the following response options (ranging from 0-3): not at all, a little, quite a bit, extremely (36). The CES-D is a 20-item questionnaire that evaluates the frequency of depressive symptoms over the past week with the following responses options (ranging from 0-3): Never or less than one day, 1-2 days, 3-4 days, 5-7 days (37). Item scores were summed to create a composite score for post-traumatic stress and depressive symptoms, respectively. The ASSIST is a comprehensive assessment used to evaluate substance use for a variety of substances such as alcohol, smoking, marijuana, and cocaine (38). The first portion of ASSIST asks about lifetime use of substances. Participants who report “Yes” for any of the substances in this portion are then asked about use of that specific substance in the prior three months (38). The first portion (e.g., any lifetime substance use) of ASSIST was used to evaluate discriminant validity. All three scales have been validated in South Africa and Zambia and are gold standard measures. Using these measures, we hypothesized that dimensions of SVAWS would be more strongly correlated with other trauma exposure measures (HTQ, i.e., convergent validity), moderately associated with depressive symptoms (CES-D) and less strongly correlated with substance use frequency (ASSIST, i.e., discriminant validity).

### Analyses

Exploratory factor analysis (EFA) with principal axis factoring (to accommodate the categorical responses to the survey questions) and promax rotation was used to evaluate the factor structure of the SVAWS (i.e., how items covaried due to underlying latent dimensions). EFA models were conducted separately in each sample to explore whether violence constructs (factors) vary across countries/cultures. Parallel analysis (e.g., comparing the eigenvalues from the data to a Monte-Carlo matrix of simulated datasets of the same size) and Scree Plots were used in tandem to determine which EFA factor solutions to evaluate for conceptual interpretability (e.g., if parallel analysis suggested a 4-factor solution, 2-, 3-, 4-, and 5-factor solutions would be evaluated (39)). Factor loading patterns were evaluated to determine how strongly each survey item was related to the underlying dimension. Items were considered for elimination if they did not appear to be related to any of the dimensions (i.e., had small nonsalient loadings (<0.3)) or appeared to be related to multiple dimensions (i.e. large crossloadings >0.3 on multiple factors). Items were iteratively eliminated, with models being reestimated after eliminating items with non-salient loadings first and items with cross-loadings second. In other words, the EFA was re-run after each item was eliminated and until there were no remaining non-salient loadings or large cross-loadings. Once the optimal factor solution was identified in each sample, Cronbach’s Alpha was calculated to evaluate internal consistency (reliability) across the subscales identified. To assess convergent and discriminant validity, the magnitude of associations was evaluated based on Pearson’s *r*.

### Ethics

The study protocol was approved by the University of the Witwatersrand Human Research Ethics Committee (Medical), the Boston University Institutional Review Board (H-39746) and the Johns Hopkins School of Public Health Institutional Review Board (12546). The study was approved as non-human subjects by the Columbia University Institutional Review Board (AAAS9661). Written informed consent was obtained from all study participants prior to enrolment by study staff for the South African study. Informed oral consent was obtained by study staff from all participants in the Zambian study as approved by the IRB. Protocol changes are approved by the IRBs and critical changes are reported to the trial registry.

## Results

### Participants

Table 1 presents baseline demographics and characteristics of both samples of women. In both countries, women were more likely to be unemployed and looking for work. In Zambia, a 39.9% were 26-35 years of age and 42.7% only completed some level of primary school. Whereas, in South Africa, 42.0% of women were 36-45 years of age and 66.8% completed primary school.

**Table 1.**
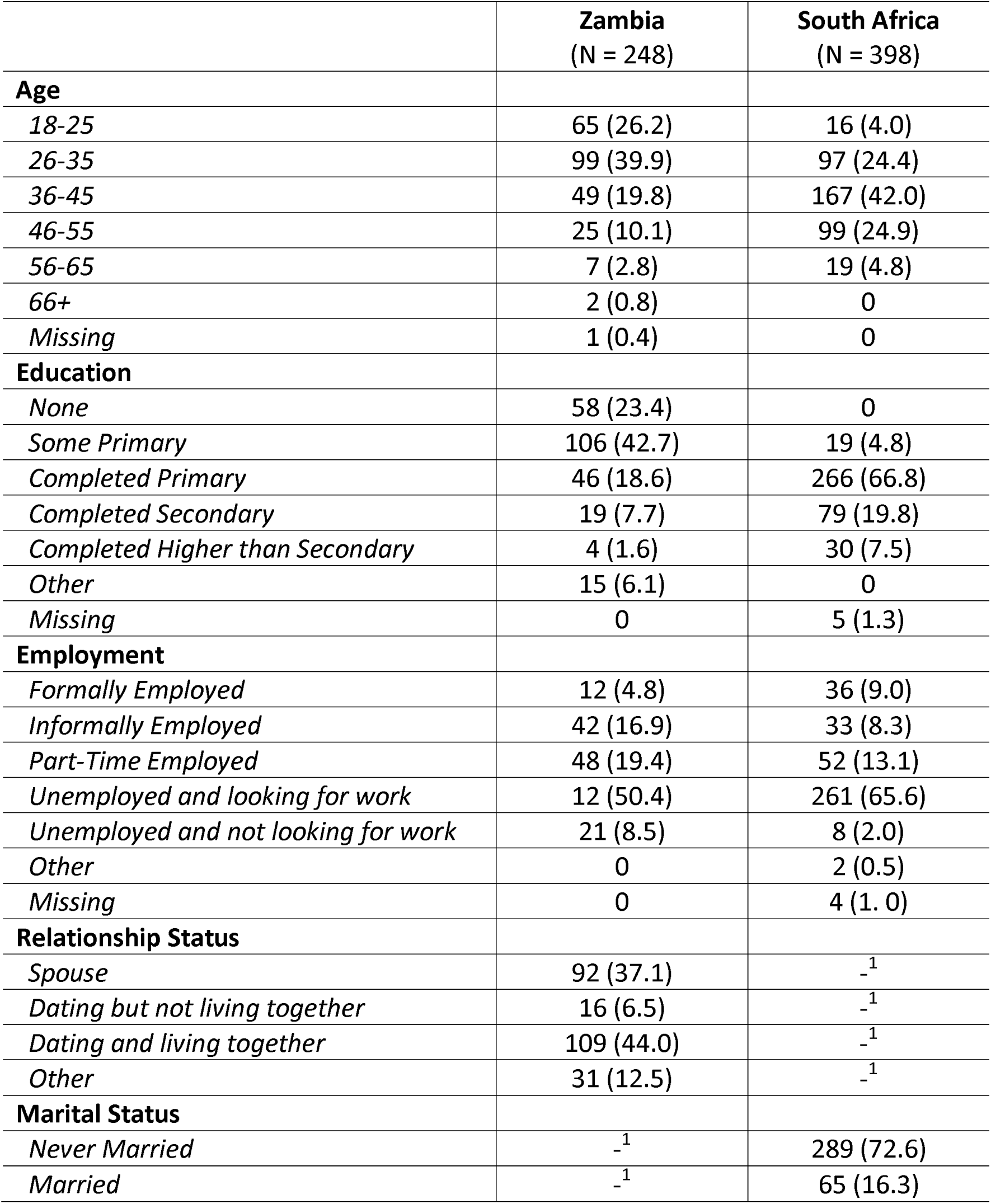
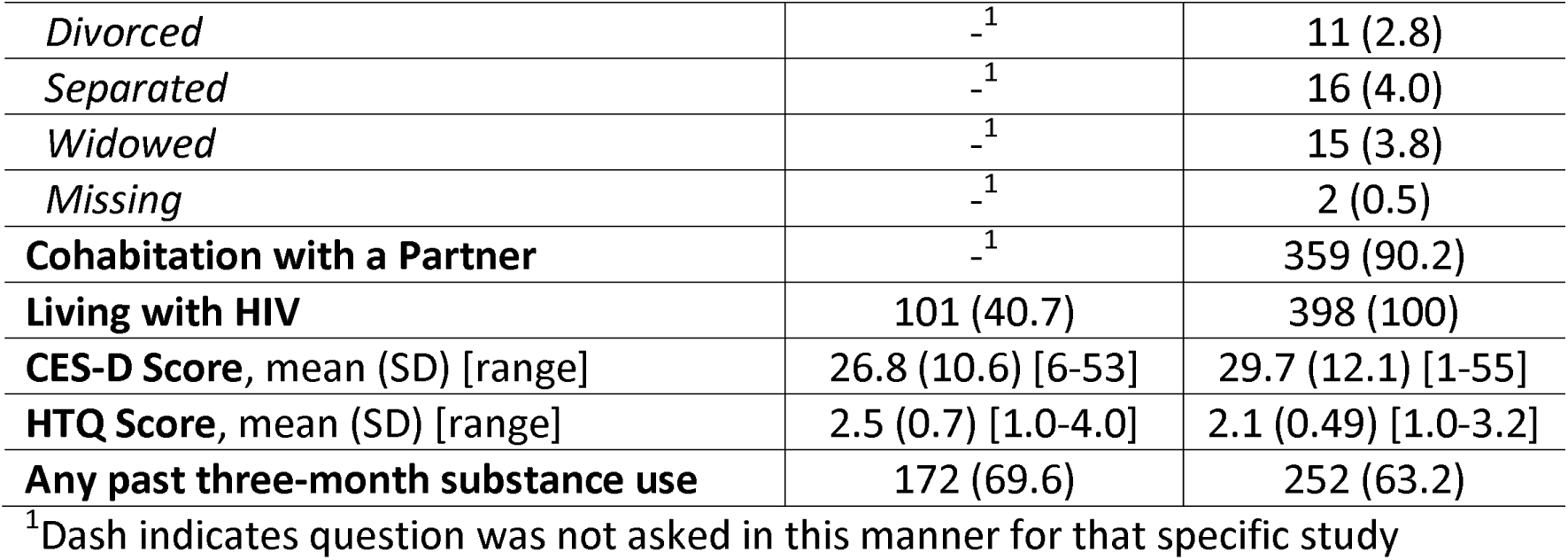
Baseline demographics for women in Zambia and South Africa.

|  | **Zambia**<br>(N = 248) | **South Africa**<br>(N = 398) |
| --- | --- | --- |
| **Age** |  |  |
| *18-25* | 65 (26.2) | 16 (4.0) |
| *26-35* | 99 (39.9) | 97 (24.4) |
| *36-45* | 49 (19.8) | 167 (42.0) |
| *46-55* | 25 (10.1) | 99 (24.9) |
| *56-65* | 7 (2.8) | 19 (4.8) |
| *66+* | 2 (0.8) | 0 |
| *Missing* | 1 (0.4) | 0 |
| **Education** |  |  |
| *None* | 58 (23.4) | 0 |
| *Some Primary* | 106 (42.7) | 19 (4.8) |
| *Completed Primary* | 46 (18.6) | 266 (66.8) |
| *Completed Secondary* | 19 (7.7) | 79 (19.8) |
| *Completed Higher than Secondary* | 4 (1.6) | 30 (7.5) |
| *Other* | 15 (6.1) | 0 |
| *Missing* | 0 | 5 (1.3) |
| **Employment** |  |  |
| *Formally Employed* | 12 (4.8) | 36 (9.0) |
| *Informally Employed* | 42 (16.9) | 33 (8.3) |
| *Part-Time Employed* | 48 (19.4) | 52 (13.1) |
| *Unemployed and looking for work* | 12 (50.4) | 261 (65.6) |
| *Unemployed and not looking for work* | 21 (8.5) | 8 (2.0) |
| *Other* | 0 | 2 (0.5) |
| *Missing* | 0 | 4 (1.0) |
| **Relationship Status** |  |  |
| *Spouse* | 92 (37.1) | _<sup>1</sup> |
| *Dating but not living together* | 16 (6.5) | _<sup>1</sup> |
| *Dating and living together* | 109 (44.0) | _<sup>1</sup> |
| *Other* | 31 (12.5) | _<sup>1</sup> |
| **Marital Status** |  |  |
| *Never Married* | _<sup>1</sup> | 289 (72.6) |
| *Married* | _<sup>1</sup> | 65 (16.3) |
| *Divorced* | <sup>-1</sup> | 11 (2.8) |
| *Separated* | <sup>-1</sup> | 16 (4.0) |
| *Widowed* | <sup>-1</sup> | 15 (3.8) |
| *Missing* | <sup>-1</sup> | 2 (0.5) |
| **Cohabitation with a Partner** | <sup>-1</sup> | 359 (90.2) |
| **Living with HIV** | 101 (40.7) | 398 (100) |
| **CES-D Score, mean (SD) [range]** | 26.8 (10.6) [6-53] | 29.7 (12.1) [1-55] |
| **HTQ Score, mean (SD) [range]** | 2.5 (0.7) [1.0-4.0] | 2.1 (0.49) [1.0-3.2] |
| **Any past three-month substance use** | 172 (69.6) | 252 (63.2) |

<sup>-1</sup>Dash indicates question was not asked in this manner for that specific study

### Determining the number of underlying factors

Scree plots generated for both the Zambian and South African samples supported a 3- or 4-factor solution (e.g., based on the “elbows” in S1 and S2 Fig.) suggesting three or four underlying violence dimensions. A parallel analysis was also conducted in each sample using 1000 simulated datasets (see dashed lines in S1 and S2 Fig. for simulated critical values). Based on these results we evaluated 2-, 3-, 4-, and 5-factor solutions. While the parallel analysis suggested a 4-factor solution would be the most ideal, the 4-factor solution as well as the 5-factor solution were found to be uninterpretable in both samples. For example, in the Zambia data the 4-factor solution was characterized by items such as breaking an object, driving dangerously with the person, beating them up, and physically forcing intercourse which assessed a combination of threatened acts, physical, and sexual violence. Of the 18 items that had loadings >0.3 onto this fourth factor, 11 cross-loaded onto other factors suggesting that this fourth factor was redundant in relation to the other three factors. A similar pattern was observed in the South African data. The 2-factor solutions were also difficult to interpret with 16 of the 22 items on the second factor also cross-loading onto the first factor. As elaborated below, the 3-factor solutions were most interpretable in both samples.

### Exploratory Factor Analysis - South Africa

Within the South African sample, 31 questions appeared strongly related to one of the three factors (e.g. had salient loadings) and the other 15 questions had either salient cross-loading (suggesting they are not clearly related to one factor) or nonsalient factor loadings (indicating that the item did not meaningfully relate to any of three factors) in the 3-factor solution (see S1 Table for factor loadings). Table 2 presents all factor loadings for items that remained after iteratively eliminating those with small loadings (<0.3) or large (>0.3) cross-loadings. The first dimension was composed of items characterized by threatening gestures *or* pointing, slapping, punching, and kicking. Slapping with the palm of the hand (0.87) and slapping the face and head (0.87) were the two items with the strongest factor loadings. These items appear to capture mild and/or indirect violence/assault without a weapon, and including threats. We labelled this as *Simple Assault.* The second dimension was comprised of items characterized by threatening with *or* using an object when physically violent, with items assessing the use of a knife or gun (0.86) and threatening with a knife/gun (0.80) having the strongest factor loadings. We labelled this as *Aggravated Threats/Assault* (i.e., severe assault with a weapon). The third dimension was composed of items characterized by demanding or physically forcing sex. Sexual intercourse against the person’s will (0.87) and demanding sex regardless of whether the person wants it or not (0.83) were the two strongest factor loadings. We labelled this as *Sexual Violence.* The three factors were moderately correlated (Pearson’s *rs* = 0.39-0.64), indicating good discrimination of the SVAWS dimensions (i.e., measuring three distinct violence subdimensions) and a lack of a hierarchical structure (i.e., second-order EFA not indicated by the data, cf. Marshall, 1992)(21).

**Table 2.**
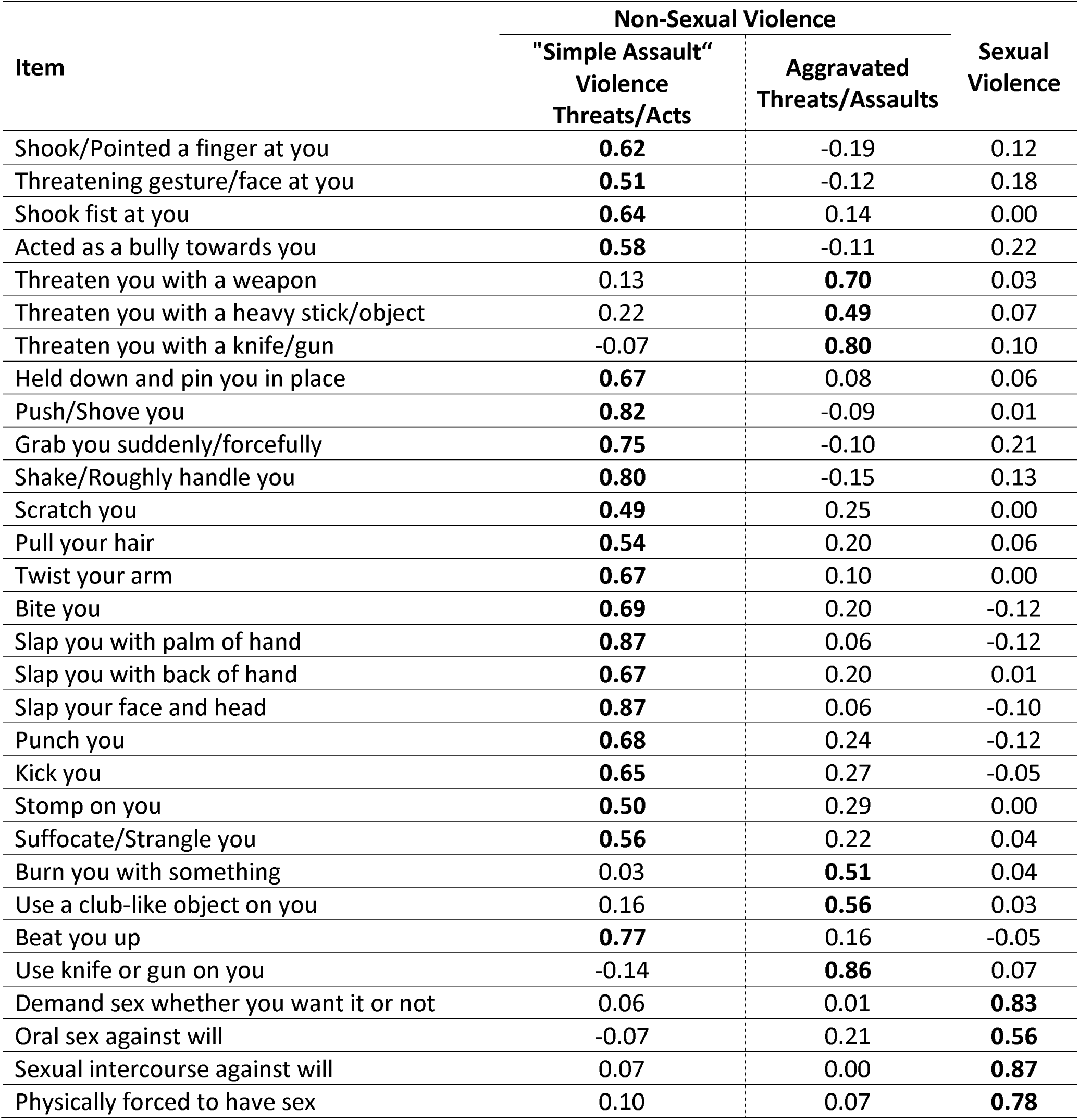
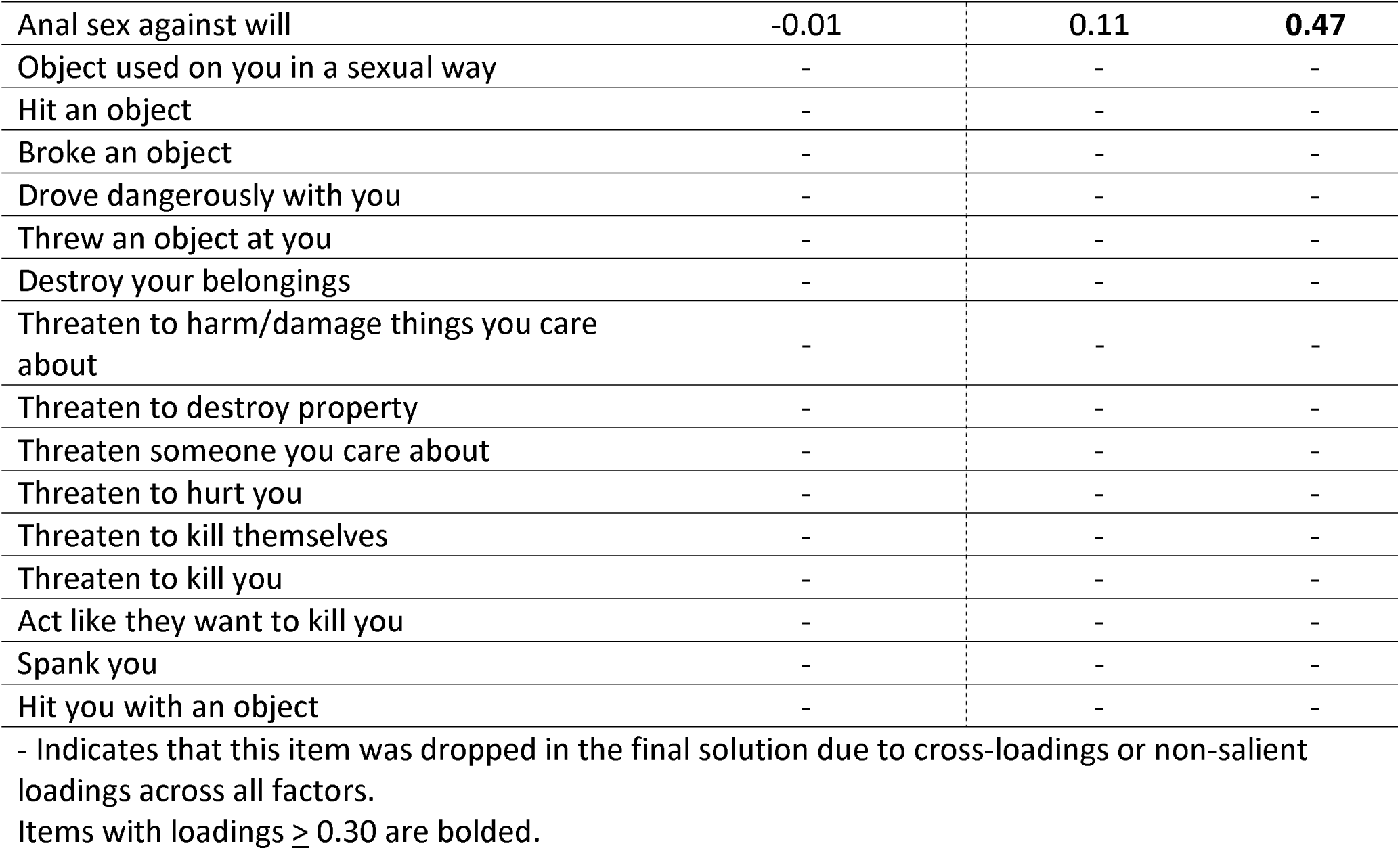
Three-factor Solution for 46-Item SVAWS in South Africa.

| Item | Non-Sexual Violence |  | Sexual Violence |
| --- | --- | --- | --- |
|  | "Simple Assault"<br>Violence<br>Threats/Acts | Aggravated<br>Threats/Assaults |  |
| Shook/Pointed a finger at you | **0.62** | -0.19 | 0.12 |
| Threatening gesture/face at you | **0.51** | -0.12 | 0.18 |
| Shook fist at you | **0.64** | 0.14 | 0.00 |
| Acted as a bully towards you | **0.58** | -0.11 | 0.22 |
| Threaten you with a weapon | 0.13 | **0.70** | 0.03 |
| Threaten you with a heavy stick/object | 0.22 | **0.49** | 0.07 |
| Threaten you with a knife/gun | -0.07 | **0.80** | 0.10 |
| Held down and pin you in place | **0.67** | 0.08 | 0.06 |
| Push/Shove you | **0.82** | -0.09 | 0.01 |
| Grab you suddenly/forcefully | **0.75** | -0.10 | 0.21 |
| Shake/Roughly handle you | **0.80** | -0.15 | 0.13 |
| Scratch you | **0.49** | 0.25 | 0.00 |
| Pull your hair | **0.54** | 0.20 | 0.06 |
| Twist your arm | **0.67** | 0.10 | 0.00 |
| Bite you | **0.69** | 0.20 | -0.12 |
| Slap you with palm of hand | **0.87** | 0.06 | -0.12 |
| Slap you with back of hand | **0.67** | 0.20 | 0.01 |
| Slap your face and head | **0.87** | 0.06 | -0.10 |
| Punch you | **0.68** | 0.24 | -0.12 |
| Kick you | **0.65** | 0.27 | -0.05 |
| Stomp on you | **0.50** | 0.29 | 0.00 |
| Suffocate/Strangle you | **0.56** | 0.22 | 0.04 |
| Burn you with something | 0.03 | **0.51** | 0.04 |
| Use a club-like object on you | 0.16 | **0.56** | 0.03 |
| Beat you up | **0.77** | 0.16 | -0.05 |
| Use knife or gun on you | -0.14 | **0.86** | 0.07 |
| Demand sex whether you want it or not | 0.06 | 0.01 | **0.83** |
| Oral sex against will | -0.07 | 0.21 | **0.56** |
| Sexual intercourse against will | 0.07 | 0.00 | **0.87** |
| Physically forced to have sex | 0.10 | 0.07 | **0.78** |
| Anal sex against will | -0.01 | 0.11 | **0.47** |
| Object used on you in a sexual way | - | - | - |
| Hit an object | - | - | - |
| Broke an object | - | - | - |
| Drove dangerously with you | - | - | - |
| Threw an object at you | - | - | - |
| Destroy your belongings | - | - | - |
| Threaten to harm/damage things you care about | - | - | - |
| Threaten to destroy property | - | - | - |
| Threaten someone you care about | - | - | - |
| Threaten to hurt you | - | - | - |
| Threaten to kill themselves | - | - | - |
| Threaten to kill you | - | - | - |
| Act like they want to kill you | - | - | - |
| Spank you | - | - | - |
| Hit you with an object | - | - | - |

- Indicates that this item was dropped in the final solution due to cross-loadings or non-salient loadings across all factors.
Items with loadings $\geq 0.30$ are bolded.

### Exploratory Factor Analysis - Zambia

Within the Zambian sample, 36 questions had salient loadings indicating they were strongly associated with one factor and the other 10 questions had either salient cross-loadings or nonsalient factor loadings in the initial 3-factor solution (see S2 Table for factor loadings). Table 3 presents all factor loadings that were not eliminated due to small loading (<0.3) or large (>0.3) cross-loadings onto multiple factors. The first dimension was comprised of items characterized by hitting, breaking, and destroying objects as well as threatening to destroy items (but not direct physical contact). The items with the strongest factor loadings were threatening gesture/face at the person (0.68) and acting as a bully towards the person (0.63). We labeled this as *Non-Contact Violence and Threats.* The second dimension was composed of items characterized by slapping, hitting with or without an object, punching, burning with *or* without an object, etc. the person. The items with the strongest factor loadings were suffocating/strangling the person (0.72) and stomping on the person (0.67). Therefore, we labelled this *Physical Violence.* The final dimension was composed of items characterized by demanding and/or forcing sex, with items assessing sexual intercourse against the person’s will (0.78) and physically forcing the person to have sex (0.78) having the strongest factor loadings. Accordingly, we labelled this as *Sexual Violence.* The three dimensions were moderately correlated (rs = 0.44-0.62), indicating good discrimination of the three dimensions and a lack of a hierarchical structure (i.e., second-order factor, cf. (21)).

**Table 3.** Three-factor Solution for 46-Item SVAWS in Zambia.

| Item | Non-Sexual Violence | Sexual |
| --- | --- | --- |

|  | **Non-Contact**<br>**Violence** | **Physical**<br>**Violence** | **Violence** |
| --- | --- | --- | --- |
| Hit an object | **0.45** | 0.04 | 0.03 |
| Broke an object | **0.62** | 0.10 | -0.08 |
| Threw an object at you | **0.49** | 0.18 | 0.00 |
| Shook/Pointed a finger at you | **0.56** | 0.05 | -0.05 |
| Threatening gesture/face at you | **0.68** | -0.15 | 0.09 |
| Shook fist at you | **0.58** | -0.04 | -0.04 |
| Acted as a bully towards you | **0.63** | -0.10 | 0.13 |
| Destroy your belongings | **0.50** | 0.10 | 0.10 |
| Threaten to harm/damage things you care about | **0.52** | 0.15 | 0.08 |
| Threaten to destroy property | **0.51** | 0.29 | -0.02 |
| Threaten to hurt you | **0.57** | 0.06 | 0.01 |
| Threaten you with a weapon | 0.06 | **0.44** | 0.21 |
| Threaten you with a knife/gun | 0.08 | **0.58** | 0.09 |
| Push/Shove you | 0.22 | **0.47** | -0.05 |
| Scratch you | 0.19 | **0.44** | -0.06 |
| Pull your hair | 0.09 | **0.40** | 0.03 |
| Twist your arm | 0.04 | **0.51** | 0.01 |
| Spank you | -0.04 | **0.56** | 0.15 |
| Bite you | -0.10 | **0.46** | 0.22 |
| Slap you with palm of hand | 0.18 | **0.51** | -0.14 |
| Slap you with back of hand | 0.10 | **0.56** | 0.06 |
| Slap your face and head | 0.19 | **0.56** | -0.10 |
| Hit you with an object | 0.00 | **0.59** | 0.07 |
| Punch you | 0.09 | **0.55** | 0.00 |
| Kick you | 0.29 | **0.45** | -0.02 |
| Stomp on you | -0.06 | **0.67** | 0.00 |
| Suffocate/Strangle you | -0.05 | **0.72** | -0.05 |
| Burn you with something | -0.21 | **0.65** | 0.11 |
| Beat you up | 0.21 | **0.46** | -0.06 |
| Use knife or gun on you | -0.04 | **0.58** | 0.11 |
| Demand sex whether you want it or not | 0.17 | -0.11 | **0.61** |
| Oral sex against will | -0.08 | 0.20 | **0.39** |
| Sexual intercourse against will | 0.13 | -0.10 | **0.78** |
| Physically forced to have sex | 0.05 | 0.03 | **0.78** |
| Anal sex against will | -0.08 | 0.13 | **0.64** |
| Object used on you in a sexual way | -0.05 | 0.20 | **0.45** |
| Drove dangerously with you | - | - | - |
| Threaten someone you care about | - | - | - |
| Threaten to kill themselves | - | - | - |
| Threaten to kill you | - | - | - |
| Threaten you with a heavy stick/object | - | - | - |
| Act like they want to kill you | - | - | - |
| Held down and pin you in place | - | - | - |
| Grab you suddenly/forcefully | - | - | - |
| Shake/Roughly handle you | - | - | - |
| Use a club-like object on you | - | - | - |

- Indicates that this item was dropped in the final solution due to cross-loadings or non-salient loadings across all factors.
Items with loadings $\geq 0.30$ are bolded.

### Internal Consistency and Concurrency Validity

Cronbach’s alphas indicated good to excellent internal reliability for all subscales in the 3-factor solution (>0.80). In South Africa, the Cronbach’s Alpha were: 0.90 for *Simple Assault,* 0.86 for *Aggravated Threats/Assault,* and 0.81 for *Sexual Violence.* In Zambia the Cronbach’s Alpha for *Non-Contact Violence and Threats* was 0.96, for *Physical Violence* was 0.87, and for *Sexual Violence* was 0.87.

Evaluation of convergent and discriminant validity indicated that all three subscales of SVAWS in both South African and Zambian samples were more strongly associated with trauma and depressive symptoms than ever reporting substance use (Table 4). For example, all 3 subscales in the South African sample were more strongly correlated with trauma *(r* > 0.33) and depressive symptoms (r > 0.34) than ever reporting substance use (r < 0.02). Similar patterns of differential magnitude of correlations were observed in the Zambian sample.

**Table 4.** Correlations *(r)* of each dimension in SVAWS with trauma (HTQ), depressive symptoms (CES-D), and lifetime substance use (ASSIST)

|  | South Africa |  |  | Zambia |  |  |
| --- | --- | --- | --- | --- | --- | --- |
|  | **“Simple Assault”**<br>**Violence**<br>**Threats/Acts** | **Aggravated**<br>**Threats/Assault** | **Sexual**<br>**Violence** | **Non-Contact**<br>**Violence** | **Physical**<br>**Violence** | **Sexual Violence** |
| Trauma | 0.45 (0.34-0.50) | 0.33 (0.23-0.41) | 0.46 (0.34-0.51) | 0.45 (0.32-0.52) | 0.49 (0.35-0.55) | 0.57 (0.41-0.60) |
| Depressive Symptoms | 0.47 (0.35-0.51) | 0.34 (0.24-0.42) | 0.36 (0.26-0.43) | 0.39 (0.25-0.47) | 0.42 (0.29-0.50) | 0.42 (0.28-0.49) |
| Lifetime Substance Use | -0.02 (-0.12, 0.09) | -0.03 (-0.13-0.07) | 0.02 (-0.08-0.12) | 0.19 (0.07-0.31) | 0.30 (0.17-0.40) | 0.29 (0.16-0.39) |

## Discussion

Results from our exploratory factor analysis identified interpretable dimensions in both samples with acceptable reliability and concurrent validity. Furthermore, results from both samples are somewhat consistent with Marshall et al.’s (1992) initial analysis of SVAWS among US women. Three of the four dimensions identified in Marshall et al,’s initial analysis are conceptually similar to the dimensions identified in the South Africa and Zambia samples. More specifically, Marshall et al. identified dimensions of threats of physical violence, actual violence, and sexual violence; the fourth dimension identified was symbolic acts (21). While our findings support the psychometric properties and utilization of the SVAWS within these two populations, several items with non-salient factor loadings or cross-loadings had to be eliminated to achieve interpretable, reliable, and valid 3-factor solutions. Our analyses also did not identify an interpretable fourth dimension as items associated with Marshall’s symbolic acts dimension were either dropped from our analysis (ex. Item 3: Drove dangerously with you in South Africa and Zambia) or were included in one of the non-sexual violence dimensions (ex. Item 1: Hit an object in Zambia). In addition, the moderate correlations among the three dimensions did not support exploration of a hierarchical factor structure (i.e., second-order dimensions, cf. Marshall et al., 1992).

These findings indicate that for SVAWS to be used in a psychometrically valid way in Zambia and South Africa, several items may need to be re-evaluated and/or considered for elimination such as “Drove dangerously with you” (Item 3). In addition, our findings suggest potential utility in defining and scoring three different IPV subscales (cf. two subscales identified by Marshall’s original validation). The use of a shorter questionnaire could improve the participant’s experience in a research study as completing a 46-item questionnaire could be time-consuming and burdensome, especially if participants need to complete additional surveys. Additional research is needed to further validate revisions to the SVAWS in these two populations exposed to high rates of IPV, especially as prior research indicates that IPV is often disclosed in a clinical setting but not formally evaluated via assessments (40).

In both samples, the sexual violence dimension appears to be similar in several ways. For instance, several of the same items loaded strongly onto the sexual violence factor (e.g. “Sexual Intercourse Against Will,” “Physically Forced To Have Sex,” “Demand Sex Whether You Want It Or Not”). Explication of a similar violence dimension across samples provides support for using the SVAWS to assess sexual IPV in both countries, in addition to suggesting similarities in the co-occurrence of sexual violence IPV behaviors (e.g., demanding and forcing sex) across the two samples and cultures and despite differences in language. In contrast, the non-sexual violence dimensions were somewhat distinct in the two samples. In South Africa, actions of mild assault/threats appear to co-occur *(”simple assault/threat” dimension)* in ways that are distinct from acts of violence with objects like sticks or weapons (*“aggravated threats/assault” dimension).* Whereas in Zambia actions of non-contact violence/threats co-occur *(“non-contact violence”* dimension} and are distinct from actions that involve physical contact/violence *(“physical assault”* dimension). These differences in non-sexual violence suggests that there is a lack of measurement invariance for SVAWS across these two cultures. Therefore, future studies may be needed to determine whether SVAWS may need to be scored and/or interpreted differently across cultures as the underlying violence dimensions being measured maybe conceptually distinct.

The conceptualization and definitions of non-sexual IPV may be different across populations, in particular these differences may be most apparent when evaluating the co-occurrence of threats versus actions and using objects versus not using objects in the context of IPV. More specifically, Zambia and South Africa may have cultural differences as to how non-sexual violence is acted out and thus defined which may explain why in South Africa the dimension of “Aggravated Threats/Assault” largely encompasses the threat and use of an object, whereas in Zambia in addition to the threat and use of an object, all forms of physical-contact violence are captured under the “Physical Violence” dimension. Prior studies have found that weapons such as guns play a significant role in violence against women in South Africa as gun ownership is highly prevalent with an estimated 1.8-million-gun owners, of whom 81% are men (41–43). Furthermore, the link between guns and increased risk of violence against women has been well established (Abrahams et al., 2010; Callaghan et al., 2024; Greenberg et al., 2024; Matzopoulos et al., 2015; Shipley et al., 2024; Small Arms Survey, 2013; Sorenson, 20172017). Additionally, there is evidence that suggests psychological abuse (e.g., Non-Contact Violence and Threats) is more prevalent than physical abuse among women in Zambia (48). This could explain why our results identified these two distinct dimensions of violence (non-contact violence/threats and physical violence) in the Zambian sample but did not in the South African sample.

Evaluating the use of SVAWS across different cultural contexts such as sub-Saharan Africa is important as we know violence and what is construed as IPV may vary across cultures. For example, prior research has found that in some communities, certain behaviors such as financial control and verbal abuse are considered disciplinary rather than abusive (29,49). Therefore, assessment of IPV may vary across different settings and can help explain why certain items in SVAWS, such as the question asking about a partner driving dangerously, are eliminated as they do not reflect aspects of violence experienced by those groups.

In these studies, SVAWS was completed by participants in several languages. In South Africa, participants were able to change their language throughout the survey therefore we were unable to determine the language in which the SVAWS was completed. While we know what language SVAWS was completed in for the Zambian sample, the number of participants in each language group was small. Therefore, we were unable to formally evaluate measurement invariance by participants’ language of response due to our small sample size overall and by language. Additionally, given our small sample size we were unable to conduct a confirmatory factor analysis and independent samples to formally test for measurement invariance. Nevertheless, the goal of this analysis was to validate the use of SVAWS in a population where several languages are spoken and evaluate whether dimensions of intimate partner violence can still be identified appropriately. Furthermore, from a feasibility standpoint, it is often not feasible for us to conduct a validation study within each language specifically within these settings given resource constraints. Despite these limitations, we were able to identify clear and consistent constructs of violence across these samples particularly the consistency of the sexual violence construct across both groups.

A revised SVAWS (dropping several items) appears to have acceptable psychometric properties and may be an appropriate tool for assessing IPV across cultures. However, given the lack of invariance demonstrated in the non-sexual violence dimensions it is important to recognize the constructs and definitions of non-sexual violence are likely different across countries and/or cultural groups. Accordingly, revision of the SVAWS or the development of new “trans-cultural” (measurement invariant) scales may be needed to assess IPV across populations. While analyses using data from the domestic violence module from the Demographic and Health Surveys indicate questions about physical IPV demonstrate measurement invariance, the data is cross-sectional and questions regarding psychological and sexual IPV lacked invariance (50). These differences emphasize the importance of evaluating surveys in new contexts to ensure concepts are measured accurately and reduce bias.

## Acknowledgements

The authors would like to thank the participants of the study.

## Data Availability

Deidentified data from the South African study will be available upon completion of the study in 2025 Deidentified data from the Zambian study are available through the SAMRC data repository: http://medat.samrc.ac.za/index.php/catalog/WW

